# Spine Reviews: Crowdsourcing Global Spine Expert Knowledge via Digital Ledger Technology

**DOI:** 10.64898/2026.04.11.26350678

**Authors:** Bassel Diebo, Guillaume Lonjon, Nassim Dehouche, Joseph Cristini, Vincent Challier, Virginie Lafage, SpineDAO

## Abstract

**Study Design:** Prospective observational study using a novel digital ledger technology (DLT)-based crowdsourcing platform.

**Objective:** To develop and evaluate “Spine Reviews,” a blockchain-based platform for aggregating spine treatment recommendations from an international specialist panel, and to validate the clinical coherence of the resulting dataset.

**Summary of Background Data:** Predictive models for low back pain treatment are limited by small, homogeneous datasets that fail to capture inter-clinician variability. Traditional multi-center data collection is expensive, slow, and geographically constrained. DLT-based crowdsourcing with cryptographic credentialing may overcome these barriers.

**Methods:** Five hundred synthetic patient vignettes (digital twins) were generated; 463 retained after quality control. A review platform was built on the Solana blockchain using non-transferable Soulbound Tokens (SBTs) for credentialing and smart-contract compensation. Fifty-two specialists from 7 countries provided ≥4 reviews per vignette across four treatment tiers, without access to imaging or physical examination. Mixed-effects regression with reviewer random intercepts partitioned decision variability.

**Results:** The platform collected 2,066 completed reviews (97.7%) over 37 days at $0.97/review. Variance decomposition revealed that 36.7% of treatment tier variability was attributable to patient presentation, 19.2% to reviewer practice style, and 44.1% to their interaction. Neurological deficits (β=0.39), symptom duration (β=0.12), and pain (β=0.09) independently predicted treatment escalation (all p<0.001). Gwet’s AC1 was almost perfect for emergency (0.92) and substantial for conservative decisions (0.67). Reviewer confidence in treatment recommendations decreased with escalating tier severity (conservative 4.59/5 vs surgical 4.05/5), suggesting appropriate uncertainty calibration.

**Conclusions:** DLT with SBT credentialing enables rapid, global, cost-effective aggregation of clinically coherent expert judgment. The three-component variance structure quantifies clinical equipoise in spine care and establishes that predictive models require diverse, multi-reviewer training data.

## Introduction

Low back pain (LBP) is the leading cause of disability worldwide and one of the most common reasons for clinical encounters across all healthcare systems.^1^ Despite its prevalence, treatment recommendations for LBP vary substantially across providers, specialties, and geographies.^2^ A patient presenting with identical symptoms may receive conservative management from one specialist, interventional treatment from another, and a surgical recommendation from a third. This variability is not random—it reflects genuine clinical equipoise: the honest disagreement among experts about optimal management for many spine conditions. Yet it creates a fundamental challenge for clinical prediction and decision support.

Artificial intelligence (AI) models for predicting LBP treatment pathways are increasingly explored as tools to reduce this variability and support clinical decision-making.^3^ However, such models are only as reliable as the data on which they are trained. Current training datasets are typically small (hundreds rather than thousands of annotated cases), geographically homogeneous (single-center or single-country), and specialty-limited (often reflecting only the surgical perspective).^3,5^ Traditional multi-center data collection—requiring institutional review board approvals, data-sharing agreements, and site coordination—is expensive, slow, and produces datasets that still reflect a narrow range of practice patterns.^4^ Critically, conventional datasets typically record the treatment decision itself without capturing the clinical reasoning, diagnostic uncertainty, or confidence level behind it. The result is AI models that may perform well within the clinical environment where they were trained but fail to generalize across settings or represent the full spectrum of expert opinion.

Digital ledger technology (DLT), commonly known as blockchain, offers a fundamentally different infrastructure for clinical data aggregation. A blockchain is a decentralized, tamper-proof record of transactions maintained across a distributed network without a central coordinating authority. Each transaction is cryptographically signed, timestamped, and permanently recorded, creating an immutable audit trail.^11,12^ In the context of medical data collection, DLT provides three capabilities not available through conventional platforms: (1) decentralized credentialing through Soulbound Tokens—non-transferable digital credentials permanently bound to an individual’s blockchain identity, encoding verified qualifications and engagement metrics;^6^ (2) automated micro-compensation through smart contracts that execute payments instantly upon task completion, eliminating institutional payment processing overhead;^12^ and (3) cryptographic provenance—an auditable chain linking every data point to a verified contributor through mathematical proof rather than institutional trust.^12^

We developed “Spine Reviews,” a DLT-based crowdsourcing platform built on the Solana blockchain that leverages these capabilities to aggregate treatment recommendations from a global panel of spine specialists. This paper describes the platform architecture, reports on its first deployment for collecting expert opinions on 500 synthetic LBP patient vignettes, and validates the clinical coherence of the resulting dataset. Rather than presenting clinical findings as primary outcomes, we use the clinical data to demonstrate that the platform produces trustworthy, internally consistent expert judgments—a necessary precondition for any downstream AI application.

## Methods

### Platform Architecture

Spine Reviews was built on the Solana blockchain, a high-performance distributed ledger processing thousands of transactions per second at minimal cost (<$0.01 per transaction). The platform comprised three integrated layers (**Figure 1**): an identity layer for reviewer credentialing, a data layer for clinical review collection and storage, and a provenance layer for on-chain payment and audit recording.

**Figure 1.**
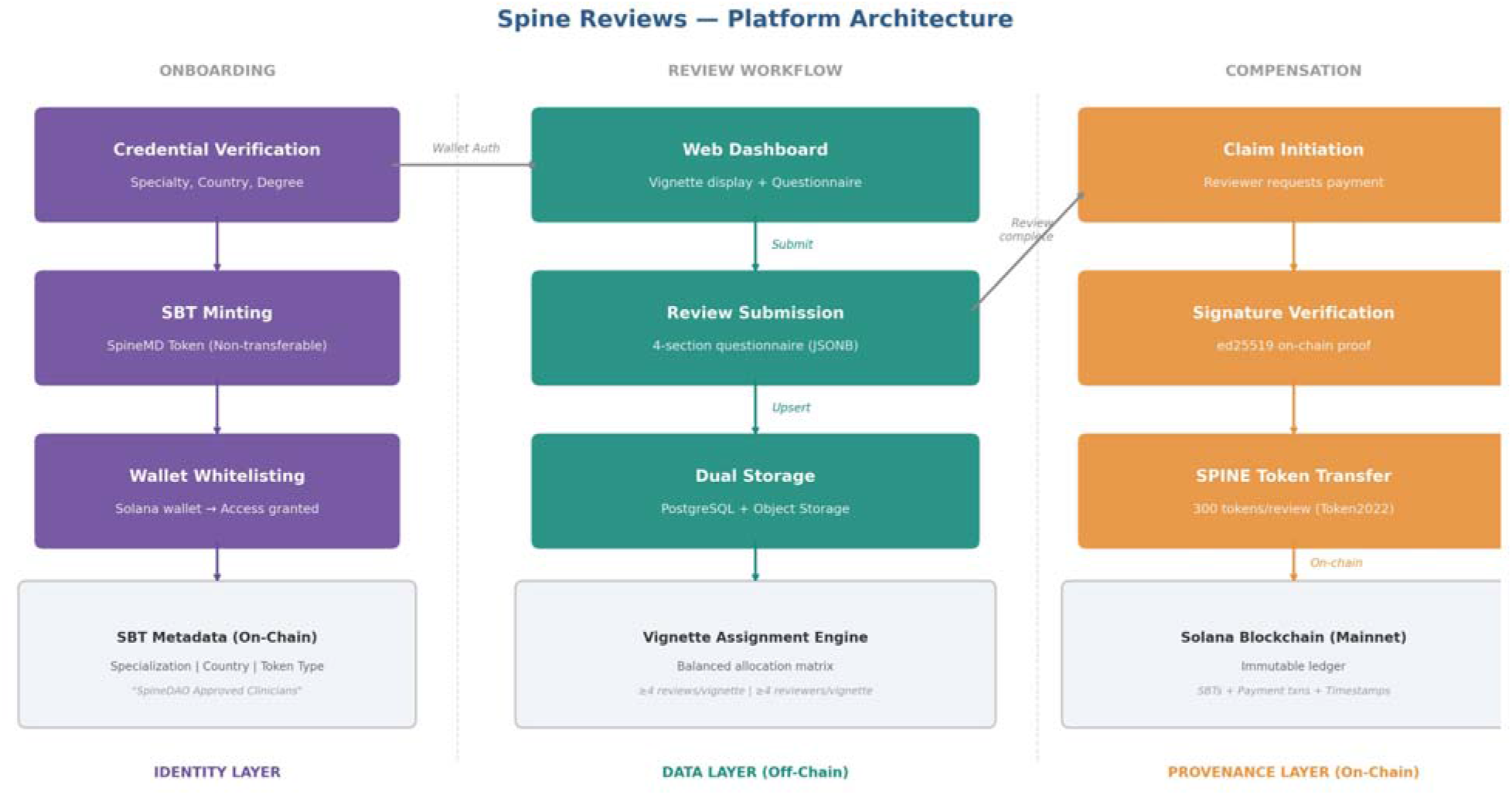
Platform architecture: identity layer (SBT credentialing), data layer (off-chain review storage), and provenance layer (on-chain payment and audit trail).

The **identity layer** was implemented using Soulbound Tokens (SBTs) minted on Solana mainnet using the Metaplex Token Metadata standard (v3) under the “SpineDAO Approved Clinicians” collection (mint address Di4Sb8jsBASeei3EJ5DD4o4qFFwrd7WkwUct3rTRyXAh). On-chain metadata (including the collection name, “SpineMD” symbol, and a URI pointing to specialization-specific JSON) were attached via a Metaplex metadata account derived deterministically from the mint address. At each identity verification event, the platform queries the wallet’s token accounts and confirms verified collection membership through the Helius Digital Asset Standard (DAS) API. Each SBT was permanently bound to a reviewer’s blockchain wallet address and encoded three on-chain attributes: medical specialization (e.g., “Orthopedic Surgery,” “Neurosurgery,” “Interventional Pain”), country of practice, and token type (“SpineMD”). Because SBTs are non-transferable by design, they guaranteed that each review was cryptographically linked to a verified specialist, no reviewer could delegate or share their credentials, and the credential record was permanently auditable on a public ledger. Credential verification was performed prior to SBT minting; the token itself then served as the persistent, tamper-proof proof of qualification.

The **data layer** comprised a Flask-based web application serving the clinical review dashboard. Reviewers authenticated by connecting their Solana wallet; the backend verified the wallet against a pre-approved whitelist and confirmed SBT ownership through the Helius Digital Asset Standard (DAS) API. Clinical review data was submitted as structured JSON and stored in a dual-redundancy architecture: PostgreSQL database (primary) with object storage backup. Critically, clinical review data was stored entirely off-chain, the blockchain recorded only identity credentials and payment transactions, not clinical content. This design preserved the benefits of blockchain provenance while avoiding the storage of potentially sensitive data on a public ledger.

The **provenance layer** handled automated compensation via Solana smart contracts. Upon review completion, reviewers initiated a payment claim through the dashboard. The backend verified: (1) the wallet’s cryptographic signature using ed25519 verification, (2) that the review was complete and unclaimed, and (3) that the wallet held a valid SBT. Upon verification, a Token2022 “transfer_checked” instruction transferred 300 SPINE tokens (approximately $0.97 USD at the time of the study) to the reviewer’s associated token account, with automatic account creation if needed. Each payment transaction was recorded on-chain with a unique transaction signature, creating a permanent, public record that links the reviewer’s identity, the review timestamp, and the compensation event.

### Patient Vignettes (Digital Twins)

Five hundred synthetic patient vignettes were generated using a Python-based pipeline (fixed random seed for reproducibility), combining clinician-curated corner cases (n=162), ensuring coverage of rare, decision-critical presentations, and constrained-random sampling (n=338) calibrated to approximate real-world clinical distributions. Thirty-seven vignettes were excluded during automated quality control for ODI computation errors, retaining 463 for reviewer assessment.

Each vignette was structured as a standardized clinical summary organized into seven domains and rendered as a two-column card (**Figure 2**):

**Figure 2.**
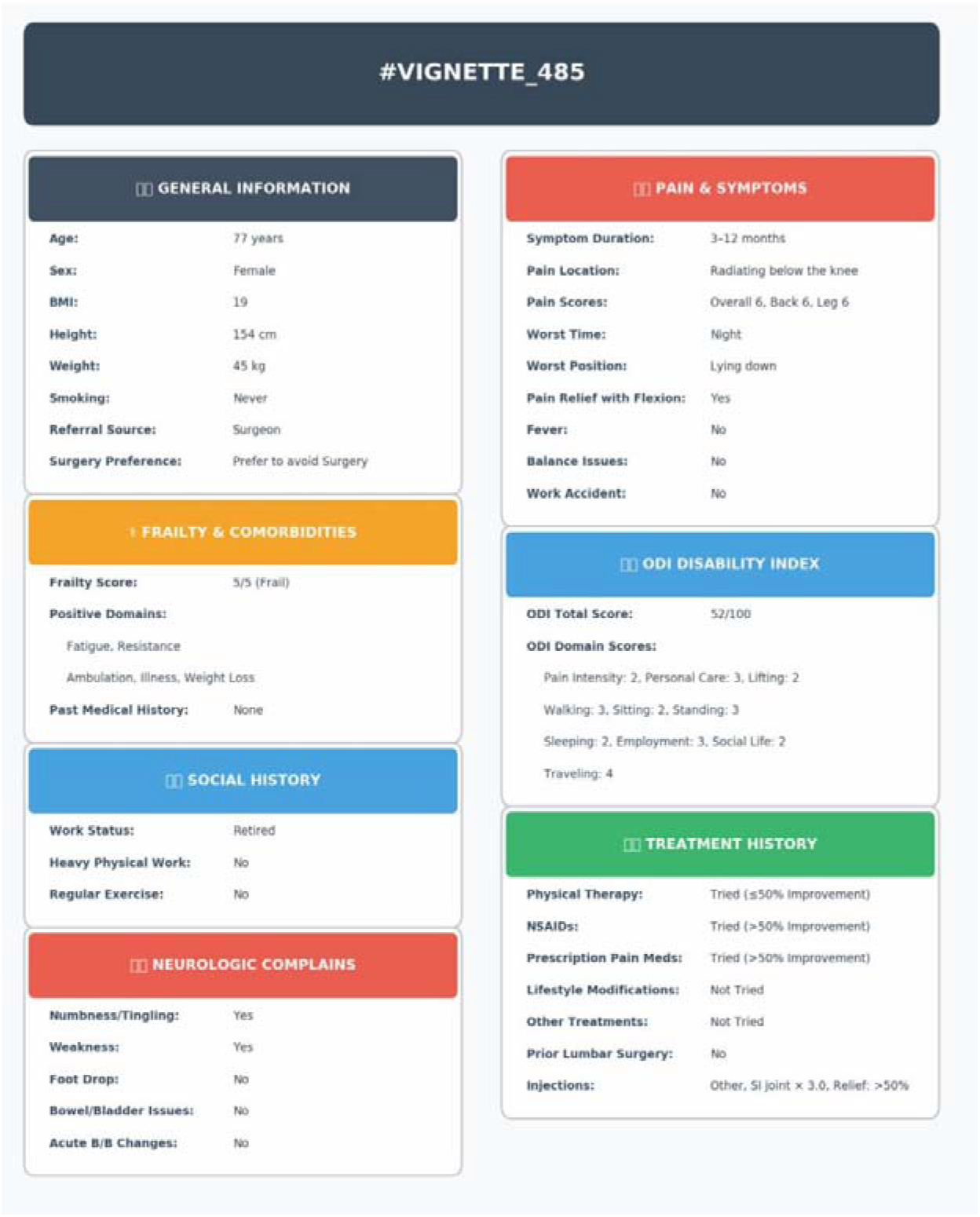
Illustrative digital twin vignettes as presented to reviewers:

- **General Information** (age, sex, height, weight, BMI, smoking status, referral source, surgery preference);
- **Frailty and Comorbidities** (FRAIL scale 0–5 with individual domains, past medical history including cancer, osteoporosis, rheumatoid arthritis);
- **Social History** (work status, heavy physical work, exercise habits);
- **Pain and Symptoms** (symptom duration in four categories, pain location, overall/back/leg pain scores on 0–10 scales, worst time and position, flexion relief, fever, balance issues, work accident);
- **Neurologic Complaints** (numbness/tingling, weakness, foot drop, bowel/bladder dysfunction, acute changes); ODI Disability Index (total score 0–100 and all ten domain scores);
- **Treatment History** (prior physical therapy, NSAIDs, prescription pain medications, lifestyle modifications, and other treatments—each graded as not tried, tried with ≤50% improvement, or tried with >50% improvement—plus prior lumbar surgery and injection history with type, number, and relief percentage).

### Review Process and Questionnaire

Fifty-two vetted spine specialists from 7 countries (France n=30, USA n=16, Switzerland n=2, Mexico, Spain, Italy, and Canada n=1 each) participated, spanning orthopaedic surgery (n=33), neurosurgery (n=10), interventional pain management (n=10), physical therapy (n=2), and rheumatology (n=1). Credentials were verified prior to SBT issuance. Vignettes were pre-assigned using a balanced allocation matrix ensuring each vignette received at least 4 independent reviews.

The review dashboard displayed the vignette card alongside a disclaimer: “Recommendations based on clinical history and patient-reported symptoms only. Physical examination findings and imaging studies (MRI, CT, X-rays) are not available for this assessment. The review questionnaire comprised three sections:

#### Section 1: Vignette Quality Check

Reviewers could exclude implausible vignettes with a coded reason (not realistic / clinical inconsistencies / missing critical information / other) and optional free-text comments, serving as a distributed quality control mechanism.

#### Section 2: Detailed Clinical Analysis

For non-excluded vignettes, reviewers assessed four treatment tiers independently (each as a binary Yes/No decision):

- **Conservative Management**: Yes/No. If yes, sub-options via checkboxes: Physical Therapy, Pain Management, Activity Modification, Bracing, Medications (NSAIDs, etc.),
- **Interventional Procedures**: Yes/No. If yes, sub-options: Epidural Steroid Injection, Facet Joint Injection, Sacroiliac Joint Injection, Nerve Ablation.
- **Surgical Consideration**: Yes/No. Free-text justification required.
- **Surgical Emergency**: Yes/No. Free-text justification required (placeholder: “If yes, explain the emergency nature: cauda equina, neurological decline, etc.”).

Critically, the four treatment tiers were independent Yes/No decisions, not mutually exclusive. Reviewers could select any combination of tiers simultaneously (e.g., conservative Yes + interventional Yes + surgical No + emergency No), reflecting the clinical reality that treatment recommendations are often staged pathways rather than single categorical choices.

#### Section 3: Additional Assessment

- **Key decision factors**: reviewers selected 1–3 factors from a structured list (ODI disability score, Pain severity, Neurological symptoms, Symptom duration, Treatment response, Age/Frailty status, Patient preference, Work/functional impact).
- **Most likely diagnosis**: free-text input. Diagnostic reasoning: free-text area for additional clinical reasoning or diagnostic considerations.

### Statistical Analysis

Treatment tiers were analyzed both as independent binary decisions and as a hierarchical outcome variable (highest recommended tier: conservative=1 < interventional=2 < surgical=3 < emergency=4). Inter-rater reliability was assessed using Gwet’s AC1, selected over Fleiss’ kappa to mitigate the kappa paradox (a well-documented phenomenon whereby extreme trait prevalence inflates chance agreement and artificially suppresses kappa values even when observed agreement is high). Binary consensus was defined as ≥75% reviewer agreement per treatment tier per vignette.

Mixed-effects linear regression with reviewer random intercepts was used for all multivariate analyses, properly accounting for the clustered data structure (multiple reviews nested within reviewers). Continuous predictors were standardized (z-scored); binary predictors were entered unstandardized. Variance decomposition partitioned total variability into between-vignette, between-reviewer, and residual/interaction components. The intraclass correlation coefficient (ICC) quantified the proportion of residual variance attributable to reviewer identity. A second model added specialty as a fixed effect to estimate how much between-reviewer variance was explained by disciplinary training versus individual practice style.

Vignette consensus phenotypes were identified using k-means clustering (k=4, selected by silhouette analysis). All analyses used Python (statsmodels, scipy, scikit-learn). No correction for multiple comparisons was applied; all p-values should be interpreted in this context.

## Results

### Platform Performance

The platform collected 2,115 reviews over 37 days, with 88% of submissions occurring within the first three weeks (**Figure 3**). Of these, 2,066 were completed (97.7%). On the 463 retained vignettes, 44 individual reviews were excluded by reviewers as clinically implausible (coded as “unrealistic” n=28, “inconsistent” n=14, “incomplete” n=7), yielding 1,917 reviews for analysis. Randomly generated vignettes were excluded at a higher rate than curated corner cases (8.9% vs 4.3%), suggesting that the constrained-random generator occasionally produced implausible combinations that clinician-curated cases avoided. Average blockchain-automated compensation was $0.97 per review. Reviewer workload averaged 36.9 reviews per specialist (SD=19.0, range 4–89), with 22 of 52 reviewers active across both halves of the collection period.

**Figure 3.**
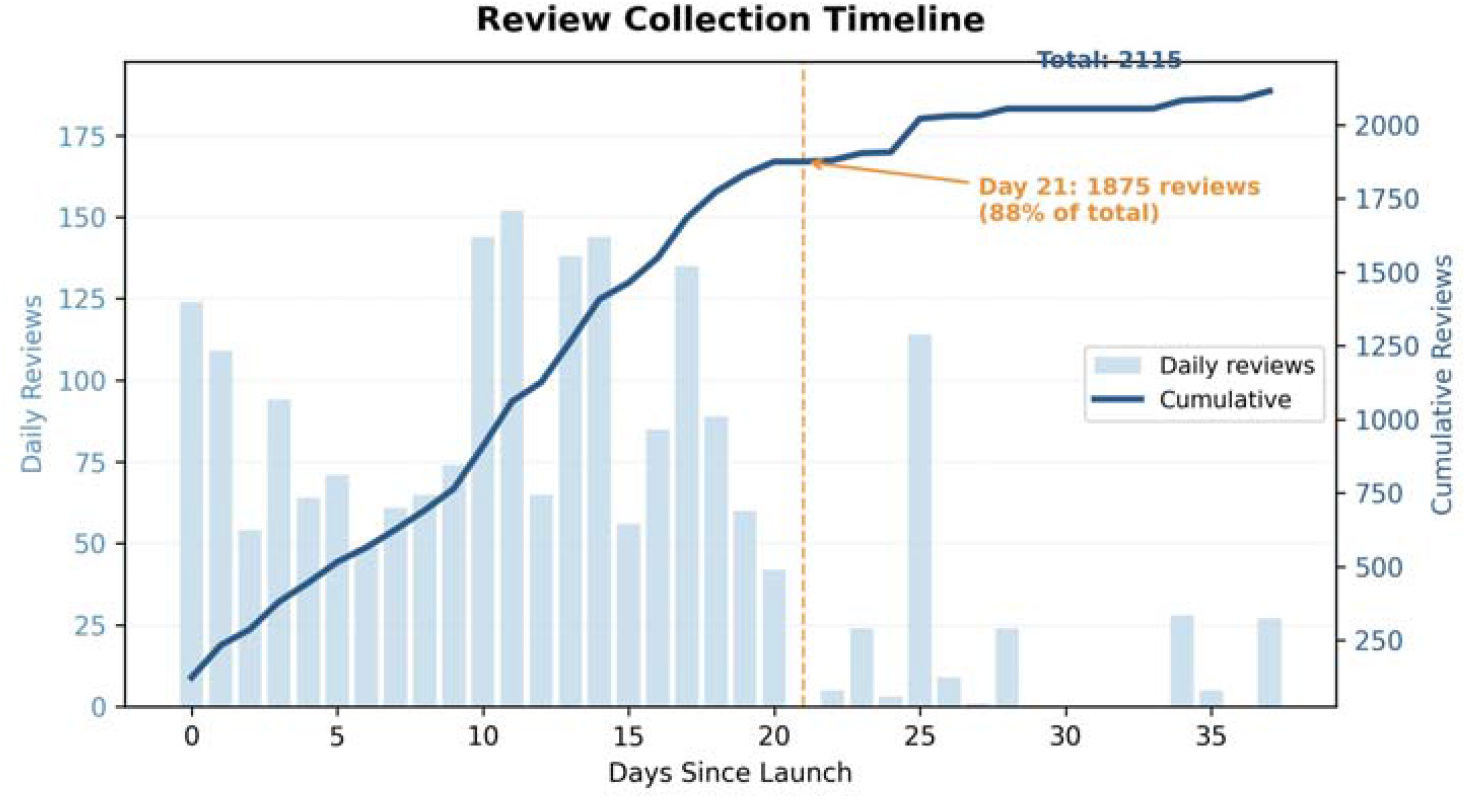
Review collection timeline. Daily submissions and cumulative total over 37 days. Dashed line marks day 21 (88% of total).

### Vignette Characteristics

Retained vignettes (n=463) are summarized in **Table 1**.

**Table 1.** Vignette characteristics. Values are n (%) or mean ± SD.

| Variable | Value (n=463) |
| --- | --- |
| Age (years), mean ± SD [range] | 52.7 ± 21.4 [18–90] |
| Sex (% male) | 51.2% |
| BMI, mean ± SD | 25.8 ± 5.2 |
| ODI total (0–100), mean ± SD | 49.6 ± 16.5 |
| Pain score overall (0–10), mean ± SD | 7.3 ± 1.3 |
| Symptom duration < 1 week | 99 (21.4%) |
| Symptom duration 1–4 weeks | 72 (15.6%) |
| Symptom duration 3–12 months | 186 (40.2%) |
| Symptom duration > 12 months | 106 (22.9%) |
| Any neurological deficit | 129 (27.9%) |
| Numbness/tingling | 111 (24.0%) |
| Weakness | 41 (8.9%) |
| Foot drop | 4 (0.9%) |
| Bowel/bladder dysfunction | 15 (3.2%) |
| Red flags present | 110 (23.8%) |
| Prior lumbar surgery | 44 (9.5%) |
| Prior injections | 161 (34.8%) |
| Vignette source: corner cases | 155 (33.5%) |
| Vignette source: random | 308 (66.5%) |

### Variance Decomposition and Clinical Coherence

The central analytical finding was the three-component variance structure of treatment decisions (**Table 2**). For treatment tier, 36.7% of variability was attributable to the vignette (patient presentation), 19.2% to the reviewer (individual practice style), and 44.1% to the residual reviewer–vignette interaction. For surgery likelihood, the pattern was similar: 41.8% vignette, 26.3% reviewer, 31.9% interaction. Neither the patient presentation nor the clinician’s identity alone can predict a treatment recommendation.

**Table 2.**
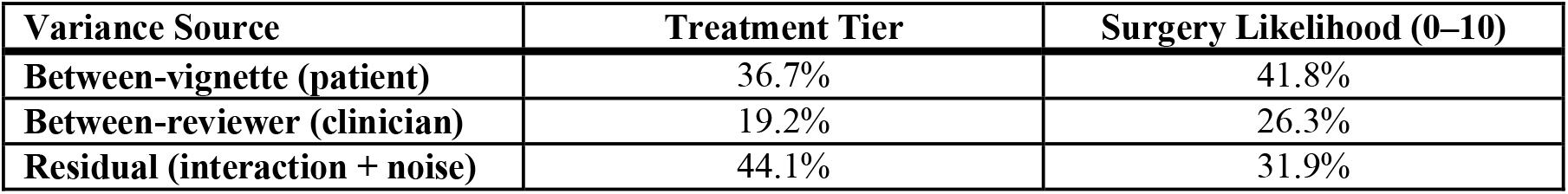
Variance decomposition of treatment decisions.

| Variance Source | Treatment Tier | Surgery Likelihood (0–10) |
| --- | --- | --- |
| Between-vignette (patient) | 36.7% | 41.8% |
| Between-reviewer (clinician) | 19.2% | 26.3% |
| Residual (interaction + noise) | 44.1% | 31.9% |

Mixed-effects regression with reviewer random intercepts confirmed that clinical severity markers independently predicted treatment escalation (Table 3). For treatment tier, neurological deficit was the strongest predictor (β=0.39, p<0.001), followed by symptom duration (β=0.12, p<0.001), pain severity (β=0.09, p<0.001), and prior injection (β=0.09, p=0.02). For surgery likelihood, neurological deficit shifted the score by over a full point (β=1.19, p<0.001), and prior surgery added 0.78 points (p<0.001). Age was a negative predictor of surgery likelihood (β=−0.13, p=0.01), reflecting risk-averse recommendations for elderly patients. ODI was a weak independent predictor (β=0.06 for tier, β=0.18 for surgery likelihood), likely reflecting the compressed ODI distribution in the synthetic vignettes (mean 49.6, only 3% <20). Red flag status was not independently predictive after controlling for neurological deficits (p>0.9).

**Table 3.** Mixed-effects regression with reviewer random intercepts. β = standardized coefficient for continuous variables, unstandardized for binary. SL: surgery likelihood (0–10). ICC: intraclass correlation coefficient.

| Variable | Tier $\beta$ | p | SL $\beta$ | p |
| --- | --- | --- | --- | --- |
| Neurological deficit | 0.392 | $<0.001$ | 1.185 | $<0.001$ |
| Symptom duration | 0.118 | $<0.001$ | 0.328 | $<0.001$ |
| Pain severity | 0.092 | $<0.001$ | 0.387 | $<0.001$ |
| Prior injection | 0.085 | 0.022 | 0.422 | $<0.001$ |
| Prior surgery | 0.107 | 0.067 | 0.781 | $<0.001$ |
| ODI score | 0.061 | 0.013 | 0.176 | 0.019 |
| Frailty score | 0.021 | 0.228 | −0.007 | 0.897 |
| Age | 0.000 | 0.990 | −0.133 | 0.014 |
| Red flag | — | $>0.9$ | — | 0.77 |
| Reviewer ICC | 0.188 |  | 0.261 |  |

Neurological findings showed graded clinical impact: mean treatment tier increased from 2.16 (no deficits) to 2.56 (any deficit), 2.83 (weakness), and 3.36 (foot drop), all p<0.001. Adding specialty as a fixed effect reduced the ICC from 0.188 to 0.155, indicating that disciplinary training explained only 18% of between-reviewer variance; the remaining 82% reflects individual practice style.

### Inter-Rater Agreement

Binary agreement per tier is reported in Table 4. Emergency AC1 was highest (0.92) but this primarily reflects universal agreement that most vignettes are not emergencies (prevalence 4.2%), not that emergency decisions are straightforward when they arise. Conservative AC1 was substantial (0.67), reflecting its role as a near-universal baseline (prevalence 83.5%), though reviewers frequently disagreed about whether to also recommend higher-intensity treatment. Interventional and surgical AC1 values were fair (0.20–0.25), reflecting genuine clinical equipoise. Importantly, AC1 captures agreement on the binary question “should this tier be included?” across all 463 vignettes; it does not capture within-vignette disagreement about treatment level, which is addressed by the consensus phenotyping analysis below.

**Table 4.**
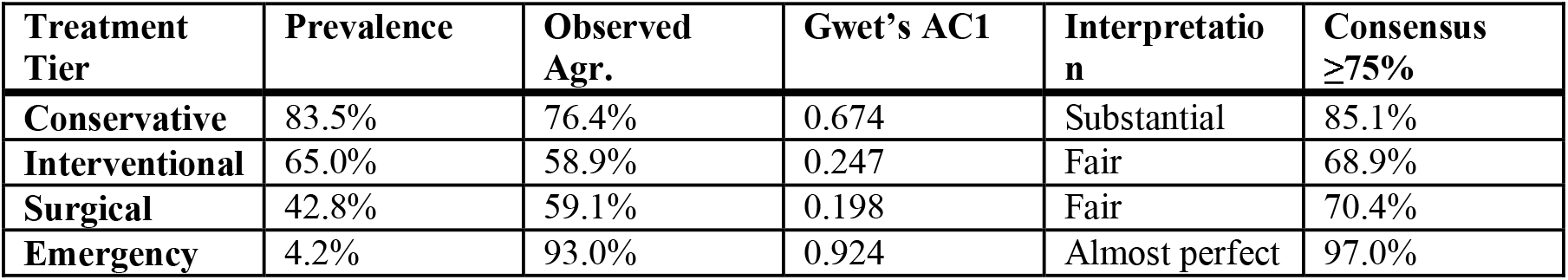
Inter-rater agreement by treatment tier. Gwet’s AC1 was used to mitigate the kappa paradox.

| Treatment Tier | Prevalence | Observed Agr. | Gwet’s AC1 | Interpretation | Consensus $\geq 75\%$ |
| --- | --- | --- | --- | --- | --- |
| Conservative | 83.5% | 76.4% | 0.674 | Substantial | 85.1% |
| Interventional | 65.0% | 58.9% | 0.247 | Fair | 68.9% |
| Surgical | 42.8% | 59.1% | 0.198 | Fair | 70.4% |
| Emergency | 4.2% | 93.0% | 0.924 | Almost perfect | 97.0% |

### Consensus Phenotyping

K-means clustering grouped vignettes by their pattern of reviewer agreement, not by treatment category (Table 5). The Surgical Convergence cluster (29%) showed the strongest consensus: 75% surgical and 75% of vignettes reaching ≥75% agreement. The Interventional Convergence cluster (37%) showed moderate consensus (41% reaching ≥75%), with 64% interventional but a quarter still recommending surgical. The Conservative-Leaning Disagreement cluster (23%) was not a consensus group: while 53% of reviews were conservative, 46% recommended interventional or surgical, and only 24% of vignettes reached agreement. The Maximum Disagreement cluster (11%) had reviews distributed across all four tiers (16%/21%/28%/32%), with only 4% reaching consensus—the same vignette receiving “conservative” from one reviewer and “emergency” from another. These clusters reconcile with Table 4: emergency AC1 is high because most vignettes are clearly not emergencies, but the 51 vignettes where emergency was considered showed the lowest consensus in the dataset.

**Table 5.** Vignette consensus phenotypes (k-means, k=4), ordered by consensus strength. Tier distribution shows % of reviews where each tier was the highest recommended (C=conservative, I=interventional, S=surgical, E=emergency)

| Cluster | n (%) | Tier Distribution (C / I / S / E) | Mean SL | Tier SD | $\geq 75\%$ Consensus | Neuro % |
| --- | --- | --- | --- | --- | --- | --- |
| **Surgical convergence** | 134 (29%) | 5 / 18 / 75 / 1 | 5.1 | 0.46 | 75% | 34% |
| **Interventional convergence** | 171 (37%) | 11 / 64 / 24 / 0 | 2.6 | 0.48 | 41% | 20% |
| **Conservative-leaning disagreement** | 107 (23%) | 53 / 22 / 24 / 1 | 2.2 | 0.73 | 24% | 14% |
| **Maximum disagreement** | 51 (11%) | 16 / 21 / 28 / 32 | 4.6 | 0.99 | 4% | 67% |

### Specialty and Geographic Variation

Orthopaedic surgeons recommended surgery in 47.7% of reviews (mean surgery likelihood 3.8/10), compared to 36.0% for neurosurgeons (3.2/10), 41.2% for interventional pain specialists (3.1/10), and 15.5% for physical therapists (1.2/10) (**Figure 4**). On shared vignettes (n=156), orthopaedic and neurosurgical recommendations were correlated (r=0.25, p=0.002) but orthopaedic surgeons recommended surgery significantly more often (46% vs 34%, paired p=0.006). Within-specialty AC1 for surgical recommendations was low for both orthopaedics (0.14) and neurosurgery (0.21), indicating most disagreement exists within, not between, specialties. Comparisons involving physical therapy (n=2 reviewers) and rheumatology (n=1 reviewer) should be interpreted with caution given underpowered samples.

**Figure 4.**
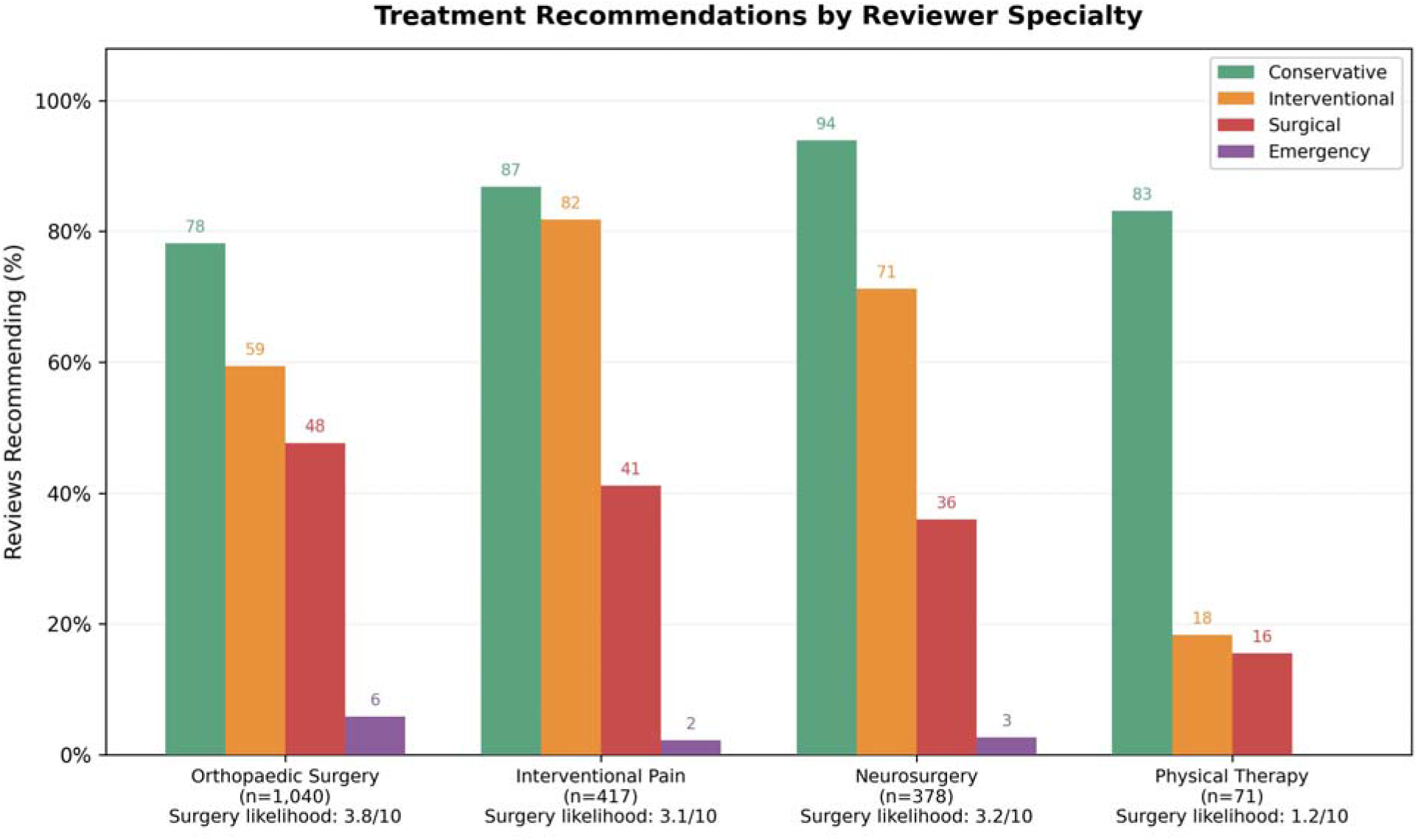
Treatment recommendations by reviewer specialty with mean surgery likelihood scores.

French reviewers (n=30, 1,048 reviews) and American reviewers (n=16, 691 reviews) showed identical surgical rates (41% vs 42%, p=0.73) and surgery likelihood (3.35 vs 3.40, p=0.75). Americans recommended interventional treatment more often (74% vs 60%, p<0.001), but after controlling for specialty and vignette features, the country effect was not significant (β=0.061, p=0.22), suggesting workforce composition rather than practice culture drove the difference.

### Treatment Escalation Patterns

The questionnaire’s multi-tier design captured the staged reasoning that characterizes real clinical decision-making: 71.3% of reviews recommended multiple treatment tiers simultaneously (mean 1.96 tiers/review). Dominant combinations were conservative + interventional (32.6%), conservative + interventional + surgical (22.1%), and conservative only (20.1%). Multi-tier recommendations were associated with higher clinical severity (ODI, neurological deficits, and symptom duration; all p<0.001).

## Discussion

This study presents Spine Reviews, a blockchain-based platform for crowdsourcing clinical expertise, and demonstrates its feasibility, efficiency, and ability to produce clinically coherent data. The platform collected over 2,000 expert reviews from 52 specialists across 7 countries in 37 days. Importantly, the data generated passed multiple internal validity checks, confirming that reviewers engaged with synthetic vignettes as they would with real patients, even in the absence of imaging studies.

### Digital Ledger Technology as Clinical Data Infrastructure

The application of DLT to medical data collection addresses three long-standing challenges. First, credentialing: in traditional crowdsourcing, verifying that a respondent holds the claimed qualifications relies on institutional trust and self-reporting. Soulbound Tokens replace this with cryptographic proof. Each SBT in the “SpineDAO Approved Clinicians” collection is a non-transferable on-chain asset encoding the reviewer’s verified specialization, country, and credential type. Because SBTs cannot be transferred, sold, or delegated, the system guarantees that every review originates from the wallet and, therefore, the individual to whom the credential was issued.

Second, compensation: traditional multi-center studies compensate reviewers through institutional payment processing, often requiring invoices and weeks of administrative delay. Smart-contract-automated payments eliminated this friction entirely, executing compensation automatically and instantaneously upon review submission with full traceability on the public ledger.

Third, provenance: the combination of SBT credentialing and on-chain payment creates an immutable audit trail linking every clinical recommendation to a verified specialist, with precise timestamping and public transaction signatures. As regulatory frameworks for AI in medicine evolve, particularly around training data transparency, cryptographic data provenance may transition from a desirable feature to a regulatory requirement.^8^

An important design decision was the separation of on-chain and off-chain data. Clinical review content was stored in PostgreSQL (off-chain), while the blockchain recorded only identity, payment, and timestamp data. This hybrid architecture preserved blockchain provenance while avoiding the impracticality and privacy concerns of storing clinical content on a public, immutable ledger.

### Clinical Coherence as Platform Validation

The clinical findings serve primarily as validation evidence for the platform. If reviewers had treated synthetic vignettes as abstract exercises, we would expect weak or inconsistent relationships between patient severity and treatment recommendations. The opposite was observed: neurological deficit, pain severity, symptom duration, and prior treatment failure all independently predicted treatment escalation.

Neurological findings showed a graded response: numbness → weakness → foot drop mapped to progressively higher treatment tiers, mirroring established surgical indications.^7^ That this coherence emerged without imaging data is noteworthy. The explicit disclaimer “Physical examination findings and imaging studies are not available” meant that reviewers relied entirely on clinical history. The finding that clinical severity markers still drove treatment escalation in the expected direction suggests that history-based vignettes capture meaningful clinical signal, though the 44% interaction variance may partly reflect reviewers’ different thresholds for recommending treatment without imaging confirmation.

The weak predictive value of ODI (β=0.06 for treatment tier) warrants discussion. This reflects the compressed ODI distribution in the synthetic vignettes (mean 49.6, only 3% below 20) rather than a genuine clinical finding. Future vignette generations should calibrate ODI distributions against clinic-level data to maximize discriminative power.

### The Variance Decomposition: Quantifying Clinical Equipoise

The three-component variance structure is arguably the most important finding for AI development. The largest component, i.e. the reviewer–vignette interaction (44%), represents how a specific clinician responds to a specific patient. This irreducible component means that neither patient severity profiles nor clinician identity can fully predict a treatment recommendation. Training data must capture diverse clinician responses to the same patients, not just diverse patients seen by a few clinicians.^3^ A model trained on five orthopedic surgeons may achieve high internal accuracy but will misrepresent the decisions of neurosurgeons, pain specialists, and even other orthopedic surgeons. Specialty explained only 18% of between-reviewer variance; the remaining 82% is attributable to individual practice style. The consensus phenotyping analysis reinforces this: 51 vignettes (11%) produced near-complete disagreement, with reviews spanning all four treatment tiers and only 4% reaching consensus—the same clinical presentation interpreted as conservative by one specialist and emergency by another.

## Data Availability

The clinical review dataset, including structured treatment recommendations and anonymized vignette responses, is available upon reasonable request to the corresponding author. The blockchain provenance records are publicly auditable on the Solana mainnet under the SpineDAO Approved Clinicians collection (Di4Sb8jsBASeei3EJ5DD4o4qFFwrd7WkwUct3rTRyXAh). The synthetic patient vignettes used in this study were generated using a validated pipeline described in Challier et al. (medRxiv 2026, doi:10.64898/2026.04.07.26350316). No clinical trial was conducted in this study. This is a platform validation study using synthetic vignettes and voluntary collection of clinician opinions. No experimental treatment was assigned to any participant. Trial registration is not applicable.

https://solscan.io/token/Di4Sb8jsBASeei3EJ5DD4o4qFFwrd7WkwUct3rTRyXAh

## Limitations

Several limitations should be acknowledged. Vignettes were synthetic and lacked imaging data, physical examination findings, and the contextual complexity of real patient encounters. While clinical coherence was confirmed, the absence of imaging represents a fundamental departure from real clinical decision-making and likely contributed to the large interaction variance component. The ODI distribution was compressed, limiting its discriminative power. The reviewer panel was concentrated in France (58%) and the USA (31%), and non-surgical specialties were insufficiently powered for robust comparisons (physical therapy n=2, rheumatology n=1). The confidence scale, while revealing a meaningful gradient across tiers, demonstrated a ceiling effect limiting its discriminative value within tiers. A slight reviewer fatigue effect was detected (mean tier drift 2.15→2.30 between first and last 10 reviews, p=0.016). Multiple comparisons were performed without formal correction, and AC1 estimates were reported without confidence intervals.

## Future Directions

The dataset—comprising structured treatment recommendations and approximately 50,000 words of bilingual free-text clinical reasoning—provides a foundation for predictive modeling, natural language processing of clinical justifications, and study of diagnostic variability (779 unique diagnostic labels, 0.4% full agreement). A critical next step is the transition from hand-crafted synthetic vignettes to vignettes generated from actual clinical data. The SpineBase multicenter spine surgery registry, currently under active data collection through the same DLT infrastructure, provides the source population for this approach. A validated synthetic data pipeline using GaussianCopula generative models with a three-domain certification framework (fidelity, utility, privacy) and blockchain-anchored provenance has been developed and benchmarked on SpineBase registry data.^10^ Applying this methodology to generate next-generation vignettes would produce digital twins that are statistically faithful to real patient distributions while preserving privacy—addressing key limitations of the current study’s manually curated vignettes, including the compressed ODI distribution. Future iterations should also expand the reviewer panel to additional geographies and non-surgical specialties, and incorporate imaging data. The platform architecture is domain-agnostic and could be adapted to crowdsourced data collection in other specialties or to peer review of scientific publications.

## Conclusions

Spine Reviews represents a first attempt to bridge Data, Human Intelligence, and AI for spine care. Digital ledger technology built on the Solana blockchain enabled rapid, global, and cost-effective aggregation of expert clinical judgment, while Soulbound Tokens created an immutable, non-transferable credibility system tracking reviewer credentials and engagement. The dataset’s clinical validity— demonstrated by appropriate treatment escalation in response to neurological deficits, pain severity, and symptom duration, even without imaging data—supports its use as a foundation for training AI models. The three-component variance structure—patient presentation (37%), clinician practice style (19%), and their interaction (44%)—quantifies the clinical equipoise inherent in spine care and establishes that meaningful predictive models require multi-disciplinary, multi-reviewer training data to capture the full complexity of treatment decision-making.

## Acknowledgements

The authors thank the 52 verified spine specialists who contributed clinical reviews through the Spine Reviews platform. Each reviewer’s credentials were cryptographically verified via Soulbound Token prior to participation; their on-chain credentials are publicly auditable on the Solana blockchain under the SpineDAO Approved Clinicians collection (Di4Sb8jsBASeei3EJ5DD4o4qFFwrd7WkwUct3rTRyXAh).

## References

1. GBD 2021 Low Back Pain Collaborators. Global, regional, and national burden of low back pain, 1990–2020, its attributable risk factors, and projections to 2050: a systematic analysis of the Global Burden of Disease Study 2021. Lancet Rheumatol. 2023;5(6):e316–e329. doi:10.1016/S2665-9913(23)00098-X

2. Lubelski D, Alentado VJ, Williams SK, Obakuwa N, Benzel EC, Mroz TE. Spine surgeon treatment variability: the impact on costs. Global Spine J. 2017;7(8):756–764. doi:10.1177/2192568217739610

3. Chou L, Ranger TA, Peiris W, Forster BB, Crofton P, Cassoobhoy M, et al. Artificial intelligence to improve back pain outcomes and lessons learnt from clinical classification approaches: three systematic reviews. npj Digit Med. 2020;3:89. doi:10.1038/s41746-020-0303-x

4. Hartvigsen J, Hancock MJ, Kongsted A, Louw Q, Ferreira ML, Genevay S, et al. What low back pain is and why we need to pay attention. Lancet. 2018;391(10137):2356–2367. doi:10.1016/S0140-6736(18)30480-X

5. D’Antoni F, Russo F, Ambrosio L, Bacco L, Vollero L, Vadalà G, et al. Artificial intelligence and computer aided diagnosis in chronic low back pain: a systematic review. Int J Environ Res Public Health. 2022;19(10):5971. doi:10.3390/ijerph19105971

6. Ohlhaver P, Weyl EG, Buterin V. Decentralized Society: Finding Web3’s Soul. SSRN Electronic Journal. 2022. doi:10.2139/ssrn.4105763

7. Kreiner DS, Hwang SW, Easa JE, Resnick DK, Baisden JL, Bess S, et al. An evidence-based clinical guideline for the diagnosis and treatment of lumbar disc herniation with radiculopathy. Spine J. 2014;14(1):180–191. doi:10.1016/j.spinee.2013.08.003

8. Regulation (EU) 2024/1689 of the European Parliament and of the Council of 13 June 2024 laying down harmonised rules on artificial intelligence (Artificial Intelligence Act). Official Journal of the European Union. 2024;L:1–144.

9. Challier V, Jacquemin C, Diebo B, Dehouche N, Denisov A, Cristini J, et al. Validated synthetic data generation from a multicenter spine surgery registry: methodology and benchmark. medRxiv. 2026. doi:10.64898/2026.04.07.26350316

10. Nakamoto S. Bitcoin: A Peer-to-Peer Electronic Cash System. 2008. Available from: https://bitcoin.org/bitcoin.pdf

11. Kuo TT, Kim HE, Ohno-Machado L. Blockchain distributed ledger technologies for biomedical and health care applications. J Am Med Inform Assoc. 2017;24(6):1211–1220. doi:10.1093/jamia/ocx068

